# Electrophysiological Correlates of Dynamic Cycling in Parkinson’s Disease

**DOI:** 10.1101/2024.02.16.24301952

**Authors:** Prajakta Joshi, Lara Shigo, Brittany Smith, Camilla W. Kilbane, Aratrik Guha, Kenneth Loparo, Angela L. Ridgel, Aasef G. Shaikh

## Abstract

**Objective:** Physical exercise like dynamic cycling has shown promise in enhancing motor function in Parkinson’s disease (PD). We examined the underlying mechanisms of dynamic cycling in PD, emphasizing its impact on the activity of the subthalamic nucleus (STN), a pivotal region within the basal ganglia.

**Methods:** The investigation involved 100 dynamic cycling sessions conducted among nine PD individuals. Each participant underwent a maximum of 12 sessions over a four-week period. Local field potentials (LFPs) originating from the STN were recorded before and after cycling, utilizing DBS electrodes positioned within the nucleus. We evaluated both immediate and sustained impacts of dynamic cycling on LFP. The periodic LFP activity was assessed by determining the dominant spectral frequency and the power associated with that frequency. Aperiodic LFP activity was analyzed by calculating the 1/f exponent of the power spectrum.

**Results:** Immediate and sustained effects of dynamic cycling on LFPs were evaluated. While immediate changes were insignificant, long-term effects showed an increasing trend in power and the 1/f exponent of the power spectrum, a measure of fluctuation in the signal, in the dorsolateral region of the STN. Ventral region of the STN did not show a significant response to the exercise intervention.

**Conclusion:** These results highlight the impact of dynamic cycling on STN neuronal activity in PD. Prolonged interventions, even without immediate changes, bring about significant modifications, emphasizing the role of extended exercise in PD management and neuroplasticity.

**Significance:** These results highlight the impact of dynamic cycling showing changing in the STN neurophysiology in PD.

**Highlights:**

1. Long-term exercise intervention affects the dorsal STN, without any immediate change, while the ventral STN shows no response.
2. Aperiodic exponent increases with exercise, mimicking the effect of dopamine
3. Periodic power also increases, behaving oppositely to its response to dopamine medication.

## Introduction

Nearly 10 million people worldwide suffer from Parkinson’s Disease (PD). The current treatment of PD is medication (levodopa, dopamine agonists) and surgical intervention (deep brain stimulation). Over the past two decades, substantial evidence has demonstrated that physical therapy incorporating various exercise methods is effective for long-term rehabilitation in PD (Mak et al., 2017; Mak and Wong-Yu, 2019). The underlying neurological mechanisms through which exercise interventions aid in PD have been studied using studies involving animal models and PD patients. Animal studies have elucidated the cellular and molecular mechanisms underlying exercise-induced neuroprotection and neurorestoration (Mak and Wong-Yu, 2019). These mechanisms include neurogenesis, neurotrophic factors, neuroinflammation, and oxidative stress (Crowley et al., 2019). Human studies have reported functional improvements in gait characteristics such as speed, stride, and step length, as well as muscle strength and balance. Additionally, enhancements in non-motor symptoms such as memory, mood, executive functions (Tiihonen et al., 2021), and overall quality of life have also been observed in response to exercise. However, in the context human studies, evidence on the mechanisms by which exercise enhances physical function is limited due to practical constraints. An fMRI study revealed a 50% improvement in following exercise, illustrating how exercise and anti-parkinsonian medication may function similarly to restore functionality (Alberts et al., 2016). Other imaging studies have also indicated that exercise interventions may induce structural changes in the brain, including increases in grey matter volume, cortico-motor excitability, striatal DA receptor density, and dopamine (DA) concentration, as observed through imaging studies. Nevertheless, the translation of these changes into modifications in neurophysiological behavior remains uncertain, particularly within the basal ganglia, where the underlying pathology of PD resides.

A previous study by (Ridgel et al., 2015) demonstrated objective improvements in motor function in PD measured with Unified Parkinson’s Disease Rating Scale (UPDRS-III) result from a structured high-cadence dynamic cycling regimen on a customized motorized stationary bike over three sessions. In this study, we aimed to understand how exercise affects the neurophysiological response of the subthalamic nucleus (STN) – a structure within the basal ganglia and a target of deep brain stimulation surgery (DBS) for PD. In participants with DBS using Medtronic Percept™ PC for a 12-session exercise regime spread over four weeks, we measured local field potential (LFP) before and after each exercise session. We aimed to understand the immediate and long-term effects of exercise by quantifying changes in the neurophysiological features of their LFP signals.

## Methods

The study protocol and written informed consent were approved by the institutional review board. The primary focus was to explore the mechanistic foundations of how dynamic cycling contributes to the improvement of PD symptoms. This involved measuring the effects of dynamic cycling on STN neuronal activity in PD. We measured effects of dynamic cycling on STN LFP activity on 100 instances from nine PD individuals (eight men and one woman; age 66.4±9.7). The average Unified Parkinson’s Disease rating scale (UPDRS-III) values were 20.7±2.56, and average daily dopamine dose was 1061.7±480 mg/day. The inclusion criteria were the diagnosis of PD confirmed with UK Brain Bank Criteria, stable antiparkinsonian medication regimen for at least six months, bilateral subthalamic region deep brain stimulation (STN-DBS) with Medtronic Percept™ PC implantable pulse generator and electrode lead models 3389 or Sense Sight B33005 Directional™ (Medtronic, Minneapolis, MN, USA), the stimulation settings being stable for at least a month’s time, the ability to provide written informed consent, and the ability to perform exercise on a motorized stationary bike every other day for up to 12 sessions over four weeks. Those with cardiovascular risk factors or musculoskeletal injury that prevents safe participation in the exercise program were excluded from the study.

The participants engaged in a structured exercise intervention that involved dynamic cycling. This intervention consisted of 12 supervised sessions spread over a duration of 4 weeks, with each session spaced 48-72 hours apart from the next. Cycling was performed on a custom made interactive motorized stationary bike at a cadence of 80 rotations per minute (RPM) for 30 mins. Each high cadence cycling was preceded by a 5-minute warm-up period and was followed by a 5-minute cool-down period at a speed of 60 RPM. The time interval between exercise and medications was kept consistent, and the participants performed the sessions at the same time of the day. Patients were advised to refrain from additional physical activities beyond their regular routine before the intervention. This precaution ensures a clear determination that any observed changes result specifically from the exercise intervention. The exercise intensity was monitored at two-minute intervals through a heart-rate monitor (Mi Band 6, Xiaomi, China or Polar H10 sensor, Polar, USA). The rating of perceived exertion (RPE) was acquired every four minutes using the 6-20 Borg RPE scale. Out of the nine participants, six completed all 12 sessions. One participant discontinued after the 9th session, and another discontinued exercise sessions after the 8th session, both due to unrelated seasonal flu like illness. One participant discontinued the study after the 11^th^ session. The discontinuation was due to unrelated circumstances. Nevertheless, we were able to test our hypothesis from all participants, who were included in the analysis.

### Exercise intervention

The intervention was done with a stationary motorized bike where the speed can be set to a desired cadence, which is 80 RPM in this case. The motor rotates the pedals at this specified speed. The rider is instructed to put in a small power to overdrive the motor. While the rider can exert additional power to pedal faster, the motor torque feedback is calibrated to maintain the set cadence. This creates a dynamic interaction between the rider and the motor. The outcome of this dynamic interaction is communicated to the rider through a visual display. The internal workings of this dynamic cycling paradigm are described in detail previously (Ridgel et al., 2017).

### LFP data acquisition

We recorded the LFP from the STN immediately before and after exercise intervention. The participants remained seated in a chair with their arms supported; they were refrained from talking or making any voluntary movements (Bougou et al., 2023; Storzer et al., 2017). The steps followed are depicted in Figure 1(a). Medication was taken at least 120 minutes before each session. During each session, the first LFP recording was obtained before starting the exercise. The DBS was turned off for 10 mins prior to beginning 210 seconds (3.5 minutes) recording at 250 Hz. The data was recorded with on-board hardware filters set at 1 Hz high-pass and 100 Hz low-pass. Following this recording, the DBS was turned back on and remained on during the biking exercise. Similar to the first recording, a second recording was obtained after completing 40 minutes of biking in the same manner as the first.

**Figure 1:**
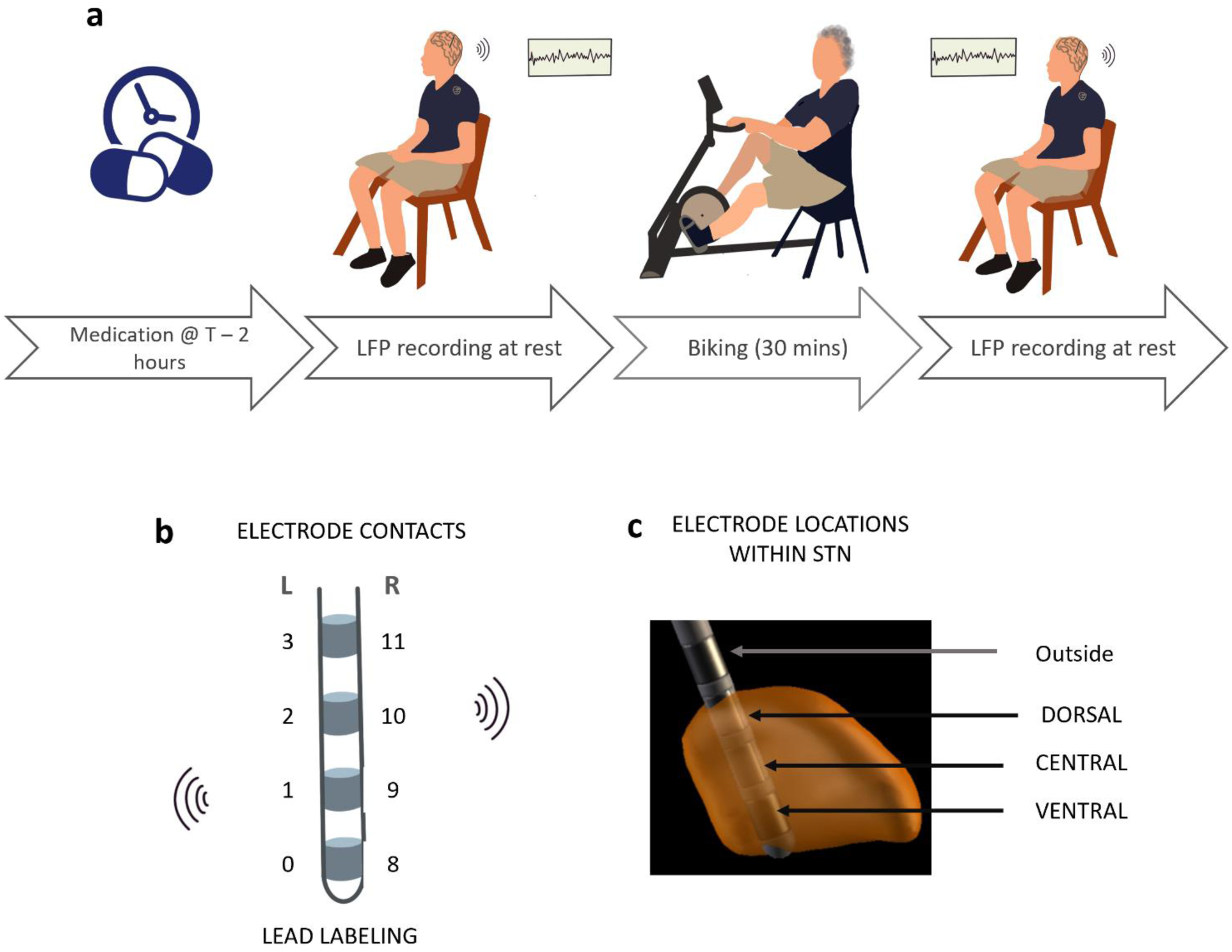
(a) shows the main steps of the experiment design protocol. The participants take medication at least 120 mins before they start exercising. We record the LFP for 210 seconds when they arrive. Later they participate in a 30 mins exercise session. At the end, we again measure the LFP signals for another 210 seconds. This process is repeated every 48-72 hours, up to 12 sessions. (b) displays the numbering of electrode contacts on DBS electrodes. Shaded areas represent electrode contacts, each labeled with a numerical designation on the left and right side, marked as L and R respectively. The left side recording involves the differential recording of 0-2 and 1-3 contacts which centered around electrodes 1 and 2 respectively. The right-side measurement comes from the differential recording from 8-10 and 9-11 contacts which are centered around electrodes 9 and 10 respectively. The Medtronic Percept™ Indefinite Streaming mode records simultaneously from these electrodes. The location of the electrodes 1,2,9,10 were identified in within the STN for each patient using Lead-DBS (Horn and Kühn, 2015). A representative of it is displayed in part (c) where the electrodes are marked as “dorsal”, “central”, “ventral”, “outside” according to their placement in the lead and used in further analysis.

We utilized the ‘indefinite streaming’ feature of the Medtronic Percept™ PC to capture LFPs. In this mode, recordings were conducted simultaneously from 6 contact pairs (3 on the right and 3 on the left) with stimulation turned off. Figure 1(B) shows the arrangement and labeling of the 4 leads on the electrode placed in each hemisphere for patients who underwent bilateral STN-DBS surgery. On the left side, the electrodes are labeled from 0 (ventral-most contact) to 3 (dorsal-most contact). Similarly, on the right side, the electrodes are labeled from 8 (ventral-most contact) to 11 (dorsal-most contact). A total of 3 differential recordings were obtained from each electrode from contact pairs (0-2, 1-3, 0-3, 8-10, 9-11, 8-11). In this study we will be only utilizing recordings from contact pairs (0-2, 1-3, 8-10, 9-11) as these pairs which provide more localized recordings than the rest.

The placement of electrodes varies among patients. To account for this variability and accurately interpret the results, it is necessary to identify the exact location of each recording pair within the STN. We used the Lead-DBS toolbox (Horn and Kühn, 2015) to reconstruct the locations of the four contact pairs of interest for each patient. We then visually classified them into three categories: dorsal, central, and ventral. Their locations were estimated based on the position of the contact situated between the two differentially recording electrodes. For instance, the location of contact pair 0-2 was assigned using the location of contact 1. Figure 1(C) shows an example of this assignment. The orange structure is the anterior view of the reconstructed right STN of PD27 using Lead-DBS. The light grey structure is the electrode, and the dark grey parts are the 4 contacts of electrode. We observed that the electrode in the lateral STN had contact 8 at the lower boundary, contact 9 in the central zone, and contact 10 in the dorsal part of the STN. From our pairs of interest, the 8-10 contact pair was designated as recording from the central region, and the 9-11 contact pair was designated as recording from the dorsal side of the STN. This process was repeated for all the contact pairs of interest in all our participants. Out of the 36 leads of interest (9 participants × 2 sides of the brain × 2 contact pairs of interest), we found 15 placed in the dorsal region of the STN, 11 in the central region, and 3 in the ventral region. The contacts found outside of the STN were not included in the analysis.

### LFP data analysis

The data was further analyzed using custom MATLAB (Version 2023b, MathWorks, United States) scripts and Fieldtrip toolbox (Oostenveld et al., 2011). After visually inspecting the electrophysiological data, any instances of movement artifacts were identified and removed from the dataset. The cardiac artifact noted in two participants was removed using the open source Perceive toolbox (https://www.github.com/neuromodulation/perceive/)(Merk et al., 2022). The output was saved for further analysis.

### Aperiodic and Periodic Component Separation

For signals obtained from each contact pair during every session, we used two methods for aperiodic component extraction: FOOOF (Donoghue et al., 2020) and IRASA (Wen and Liu, 2016). For FOOOF, the LFP data was filtered using a 5th order, zero phase Butterworth filter from 4 Hz to 60 Hz. The power spectral density (PSD) was calculated using Welch’s method. This data was then processed with the FOOOF algorithm with the following parameters: frequency range of 8-50 Hz, peak width limits of 2-20 Hz, a maximum of 6 peaks, a minimum peak height of 0, a peak threshold of 2, and an aperiodic model set to fixed. After computing the aperiodic fit, we subtracted the aperiodic values from the full PSD to obtain the periodic PSD and recorded the aperiodic exponent values. Figure 1 illustrates this process. The recorded signal (Figure 2A) was filtered with a 4-60 Hz filter, and the PSD (Figure 2B) was computed. The PSD was then separated into aperiodic – depicted by blue curve in (Figure 2C) and periodic components (Figure 2D) using the FOOOF algorithm.

**Figure 2:**
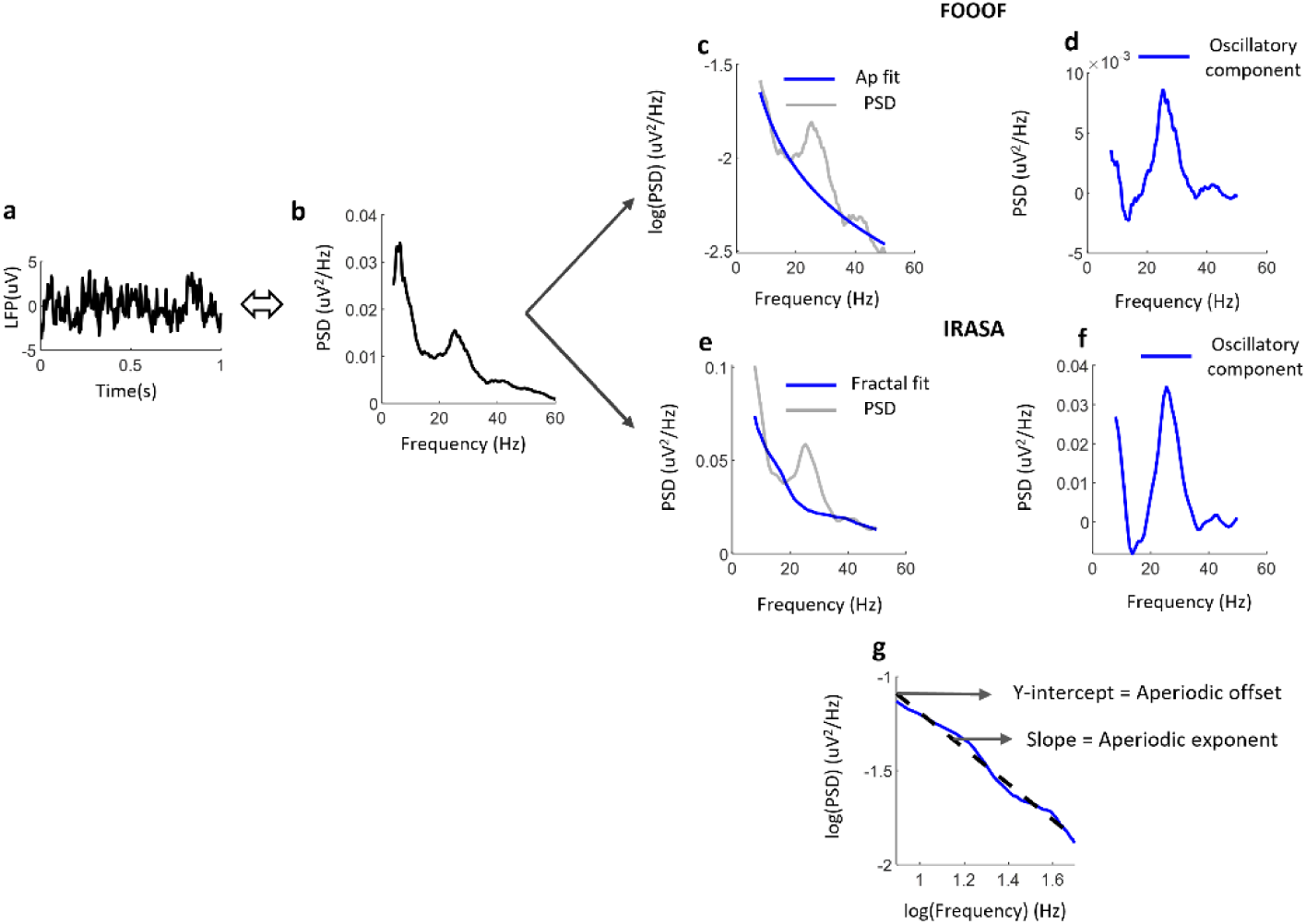
(a) displays a 1-second epoch of a sample LFP signal from one participant. (b) illustrates the power spectral density curve of the entire signal that includes the 1-second epoch shown in (a). (c) presents the aperiodic spectrum (blue) computed using the FOOOF algorithm, with the grey curve representing the raw PSD used for the fit. (d) shows the oscillatory component remaining after removing the aperiodic fit. Similarly, (e & f) shows the aperiodic fractal component calculated using the IRASA algorithm. (e) is the fractal fit, and (f) is the oscillatory component.

For IRASA, each dataset was then divided into 1-second windows with 50% overlap. We used the default resampling factor, hset = (1.1 to 1.9 in increments of 0.05), and a frequency range of 8-50 Hz to compute the aperiodic spectrum (Figure 2E), without applying any digital filters to the data beforehand. The resulting fractal curve was subtracted from the original PSD to derive the periodic spectrum (Figure 2F). The aperiodic exponent and aperiodic offset were determined by calculating the slope of the linear fit of the aperiodic curve in the log (PSD)-log (frequency) domain (Figure 2G). Due to the imperfections of both algorithms as described in (Gerster et al., 2022), we compared the results from both algorithms to validate our outcomes.

In addition to the aperiodic exponent derived from the aperiodic fit, two features were extracted from the periodic traces obtained using both methods: peak power and the frequency at which this peak power occurred. Peak power represents the maximum power detected within the 8 to 50 Hz range, with the corresponding frequency identified as the “frequency corresponding to the peak power”.

### Statistical analysis

To compare data parameters before and after biking in the acute analysis, we used the independent non-parametric Wilcoxon signed rank test as the data did not follow a normal distribution, as determined by the Kolmogorov-Smirnov test. Additionally, Spearman’s rank correlation analysis was performed to examine the relationship between session number and LFP characteristics across median values of dorsal, central, and ventral contacts, aiming to identify any significant monotonic trends in each feature.

## Results

The overarching goal of this study was to gain insights into the physiological underpinning of how a short-term and long-term dynamic cycling intervention affects PD, particularly in the context of STN neuronal activity. We initially hypothesized that changes in LFP features would be observed both acutely and over the long term. To examine this hypothesis, we studied LFP signals collected from before and after the biking paradigm from 9 PD participants with bilateral STN DBS using the Medtronic Percept™ PC device.

### Immediate effects of dynamic biking on STN activity

We analyzed LFP data from a total of 100 sessions from 9 participants immediately before and after biking. The comparison of LFP features, including the aperiodic exponent, peak power, and the frequency corresponding to the peak power, did not show any significant changes in the dorsal, central, and ventral parts of the STN. Figure 3 (A-I) summarizes these findings. In each plot, the X-axis represents the value of each LFP feature before biking, and the Y-axis represents the value of the same feature on the same day after biking. The broken black line indicates the 95% confidence interval for the data, and the solid black line (x=y) passes through the region within this interval.

**Figure 3:**
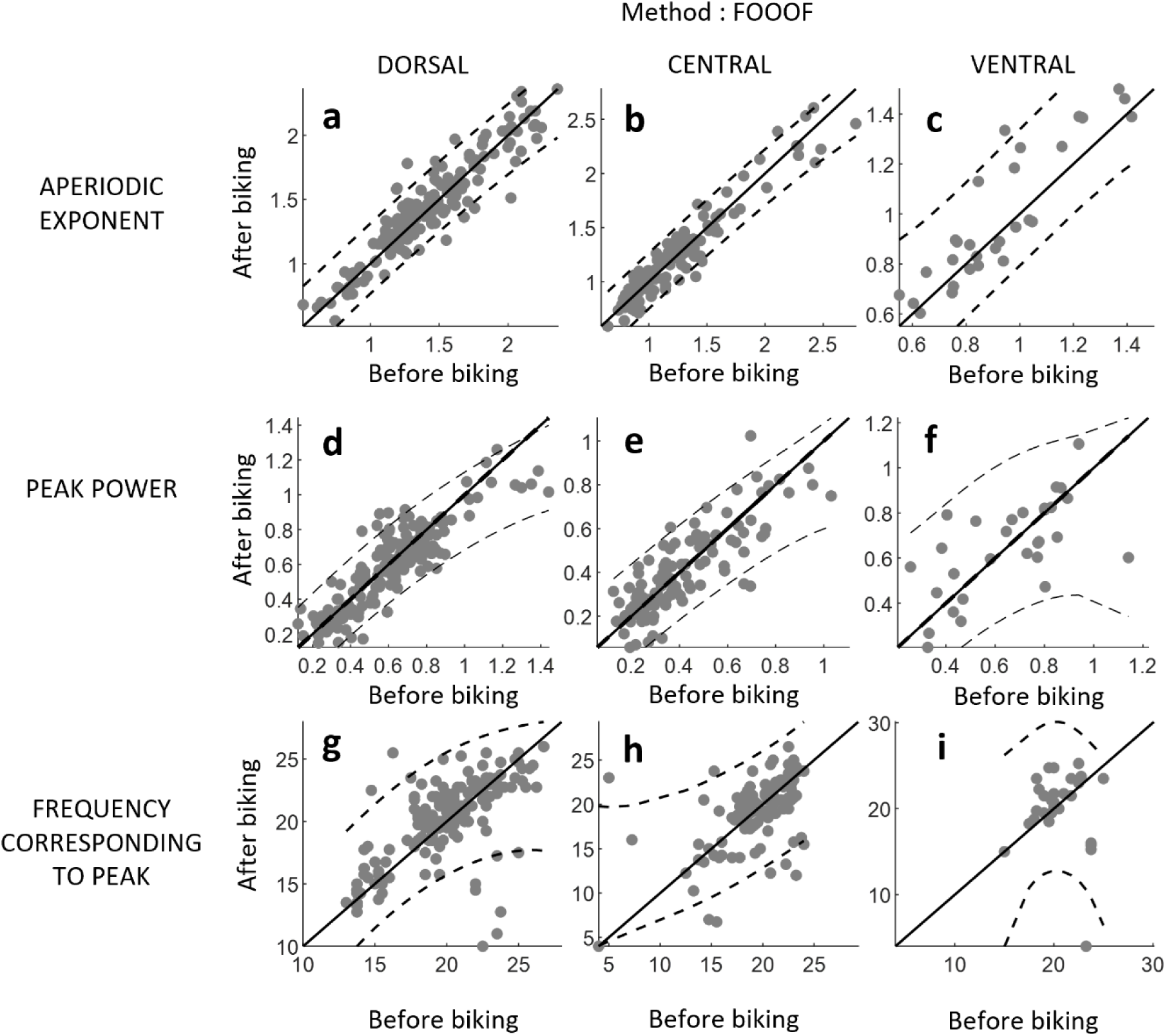
(a-i) summarizes the findings showing the acute response of exercise on the STN LFP parameters calculated using the FOOOF algorithm. In each plot, the X-axis represents the value of each LFP feature before biking, and the Y-axis represents the value of the same feature on the same day after biking. The broken black line indicates the 95% confidence interval for the data, and the solid black line (x=y) passes through the region within this interval. (a, b, c) show the values of aperiodic exponents in the electrodes placed in the dorsal, central, and ventral regions, respectively. (d, e, f) show the value of the peak power detected within the frequency range 8-50 Hz in the dorsal, central, and ventral regions. (g, h, i) show the values of the frequencies corresponding to the peak power in the dorsal, central, and ventral regions.

When performing a paired comparison using the Wilcoxon signed rank test on the aperiodic exponents of the dorsally placed electrodes, the results showed a p-value of 0.06. Before biking, the median was 1.37 with an interquartile range (IQR) of 1.22 to 1.62. After biking, median was 1.41 with an IQR of 1.22 to 1.64. Similarly, the central electrodes exhibited a p-value of 0.83, with median values of 1.17 (IQR: 0.95-1.4) before biking and 1.20 (IQR: 0.98-1.38) after biking. In contrast, the comparison in ventral electrodes showed a p-value of 0.02, with median values of 0.9 (IQR: 0.76-1.04) before biking and 0.88 (IQR: 0.79-1.2) after biking. Despite the statistical significance observed, the change in median values is 2.2%. The peak power showed a significant change in the dorsal electrodes, with a p-value of 0.02 and a median of 0.62 (IQR: 0.45-0.75) before biking, compared to 0.62 (IQR: 0.41-0.74) after biking, indicating a change of less than 1% in the median value. In contrast, the central and ventral locations showed no significant changes, with p-values of 0.47 and 0.94, respectively. The median for the central location was 0.35 (IQR: 0.26-0.50) before biking and 0.35 (IQR: 0.27-0.53) after biking, while for the ventral location, it was 0.71 (IQR: 0.43-0.80) before biking and 0.66 (IQR: 0.52-0.80) after biking. The frequency at which peak power occurred also demonstrated similar trends. Significant changes were observed on the dorsal side (p-value 0.01), with a median of 20.25 (IQR: 18.5-22.5) before biking and 21 (IQR: 18.5-22.7) after biking, reflecting a magnitude of change of 3.7%. In contrast, the central and ventral locations did not exhibit significant changes. The centrally placed electrodes had a change with a p-value of 0.64 and medians of 19.9 (IQR: 18.25-22.3) before exercising and 20 (IQR: 17.6-21.8) after exercising. The ventrally placed electrodes showed a change with a p-value of 0.17 and median values of 20 (IQR: 19-22.6) before biking and 21 (IQR: 18.7-23.5) after biking, respectively.

### Long-term effects of dynamic cycling on STN activity

To understand the long-term effects on STN activity, we analyzed LFP data collected from each participant before their biking exercise over 12 dynamic cycling sessions performed over 4-week period. The data was consistently collected at the same time of day and the same duration after they took their medication. We calculated the percentage change in each LFP feature (aperiodic exponent, peak power, and frequency corresponding to the peak power) relative to its baseline value from session number 1. Figure 4(A-I) illustrates the distribution of these values across sessions. Each error bar entry displays the median value for each session across all electrodes positioned in that location, with the value for that session represented by a solid black circle and the interquartile range shown by grey error bars. The dashed blue line indicates the linear fit through the medians. The “rho” value represents the Spearman correlation coefficient calculated between the session count and the medians. The “p” value indicates the statistical significance of this Spearman correlation. We found significant trends in LFP feature changes among dorsally placed leads, whereas central and ventral leads did not exhibit notable trends.

**Figure 4:**
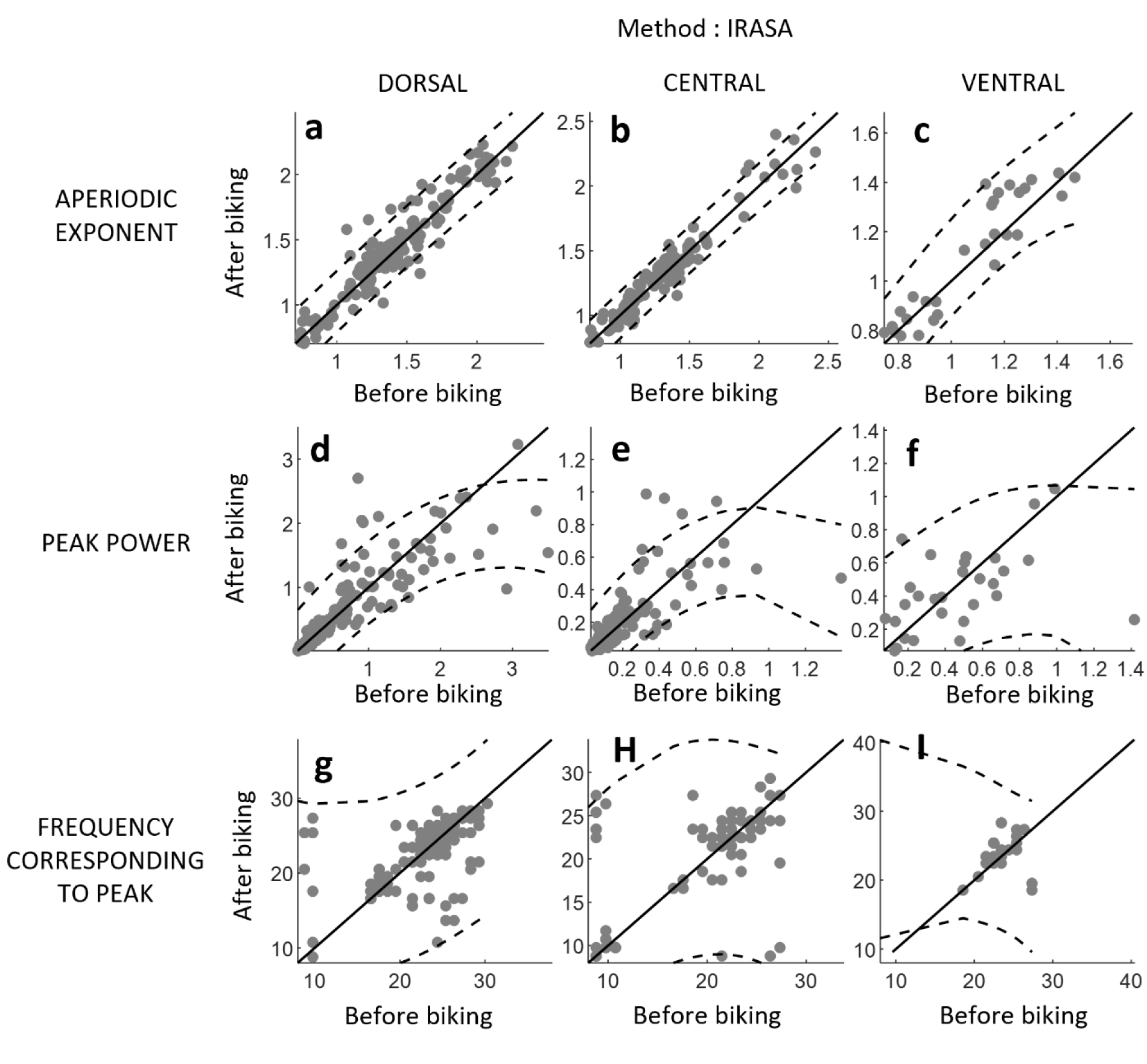
(a-i) summarizes the findings showing the acute response of exercise on the STN LFP parameters calculated using the IRASA algorithm. Each plot has the X-axis representing the value of each LFP feature before biking and the Y-axis representing the value of the same feature on the same day after biking. The broken black line indicates the 95% confidence interval for the data, while the solid black line (x=y) passes through this interval. (a, b, c) display the values of aperiodic exponents in electrodes placed in the dorsal, central, and ventral regions, respectively. (d, e, f) show the values of peak power detected within the frequency range of 8-50 Hz in the dorsal, central, and ventral regions. (g, h, i) illustrate the frequencies corresponding to the peak power in the dorsal, central, and ventral regions.

For example, dorsally placed leads showed a significant increasing trend (Figure 4A) (rho = 0.62, p = 0.04). In contrast, central (p = 0.68) (Figure 4B) and ventral leads (p = 0.94) (Figure 4C) did not exhibit significant trends. This suggests that dorsal leads responded with an increasing aperiodic exponent over sessions. A similar trend was observed for peak power, which showed a progressive increase across sessions, as depicted in Figure 4D. The medians indicated a Spearman correlation (rho = 0.64, and p = 0.03), indicating an increasing trend in peak power with the biking intervention. The frequency corresponding also showed a significant trend in the dorsal leads but did not show any significant correlation in the central (p = 0.51) and ventral (p = 0.23) leads. The frequency corresponding to peak power (Figure 4G) also demonstrated significant trends in dorsally placed leads but not in centrally and ventrally placed leads. Specifically, the frequency showed a decreasing trend over sessions (rho = −0.65, p = 0.02) in dorsal leads, whereas central and ventral leads showed non-significant p-values of (p = 0.51) and (p = 0.23), respectively. This suggests a slowing down of the frequency at which neuronal oscillators are active in the dorsal region. Overall, there is a consistent response observed in dorsal leads, while centrally and ventrally placed leads do not show consistent trends. This indicates that only the dorso-lateral part of the STN responded to this exercise intervention.

We conducted the same analysis using the IRASA algorithm. The results closely mirrored those obtained previously, with minor variations in values but consistent conclusions drawn from the statistical analysis. Figure 5 (A-I) similarly reflects results when calculating the aperiodic exponent using the IRASA algorithm. Despite the equality line passing through the confidence intervals, no significant changes were observed for any of the features across electrode locations (p-values > 0.05). Furthermore, results obtained using FOOOF, which indicated significant p-values, revealed minimal changes in the median (<4%). Thus, we conclude that the STN-LFP features did not exhibit substantial alterations immediately in response to biking.

**Figure 5:**
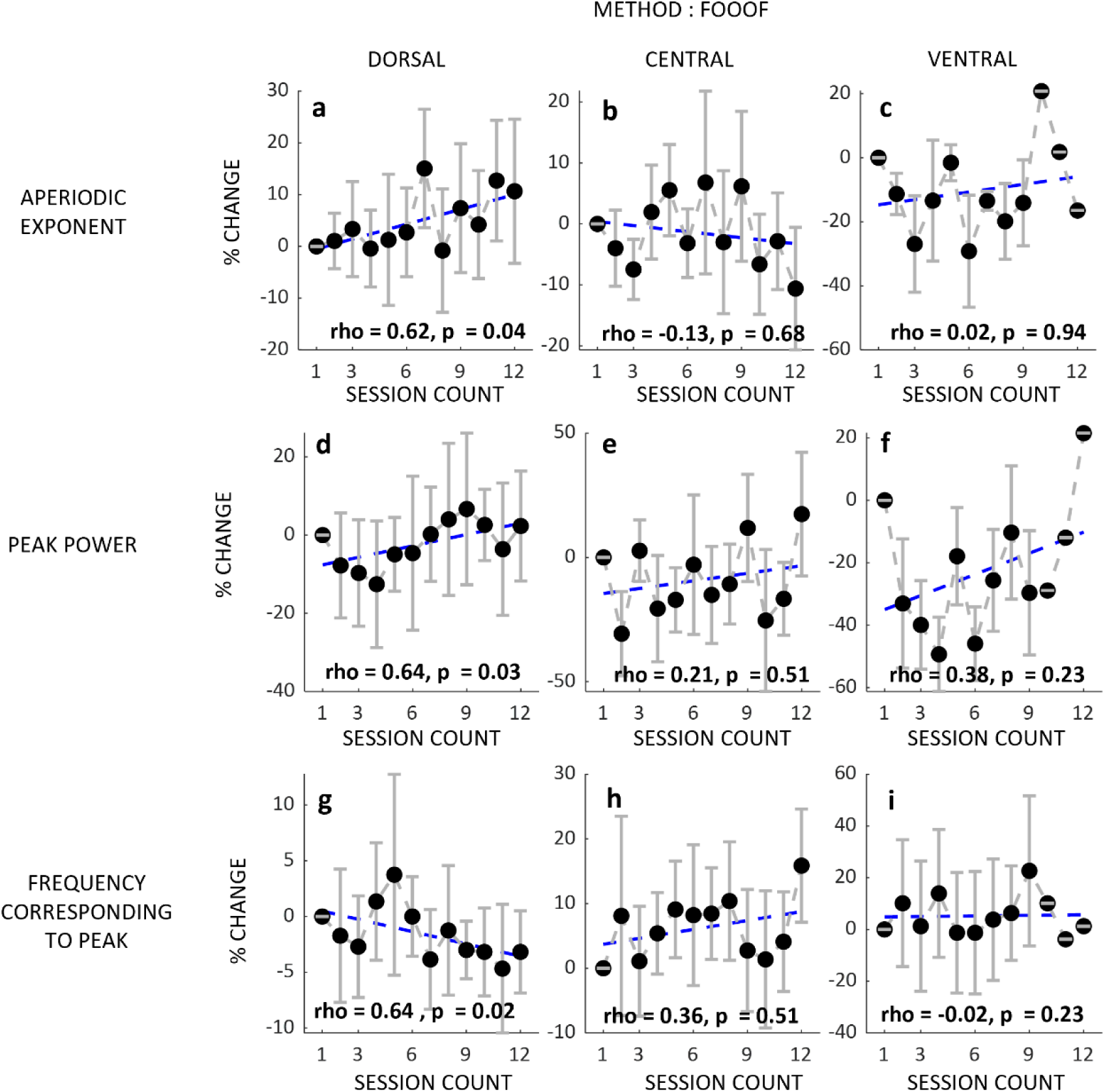
(a-i) illustrate the trend in the change in LFP features throughout the 12 exercise sessions. For each electrode placed dorsally, centrally, and ventrally, the percentage change from the respective baseline was calculated using the FOOOF algorithm. The median value for all electrodes for each session is marked with a filled black circle, and the light grey error bars show the interquartile range. (a-c) depict the change in the aperiodic exponent in the dorsal, central, and ventral regions, respectively. (d-f) show the incremental changes in peak power detected within the 8-50 Hz frequency range, and (g-i) display the changes in the frequency corresponding to the peak values as the sessions progress.

**Figure 6:**
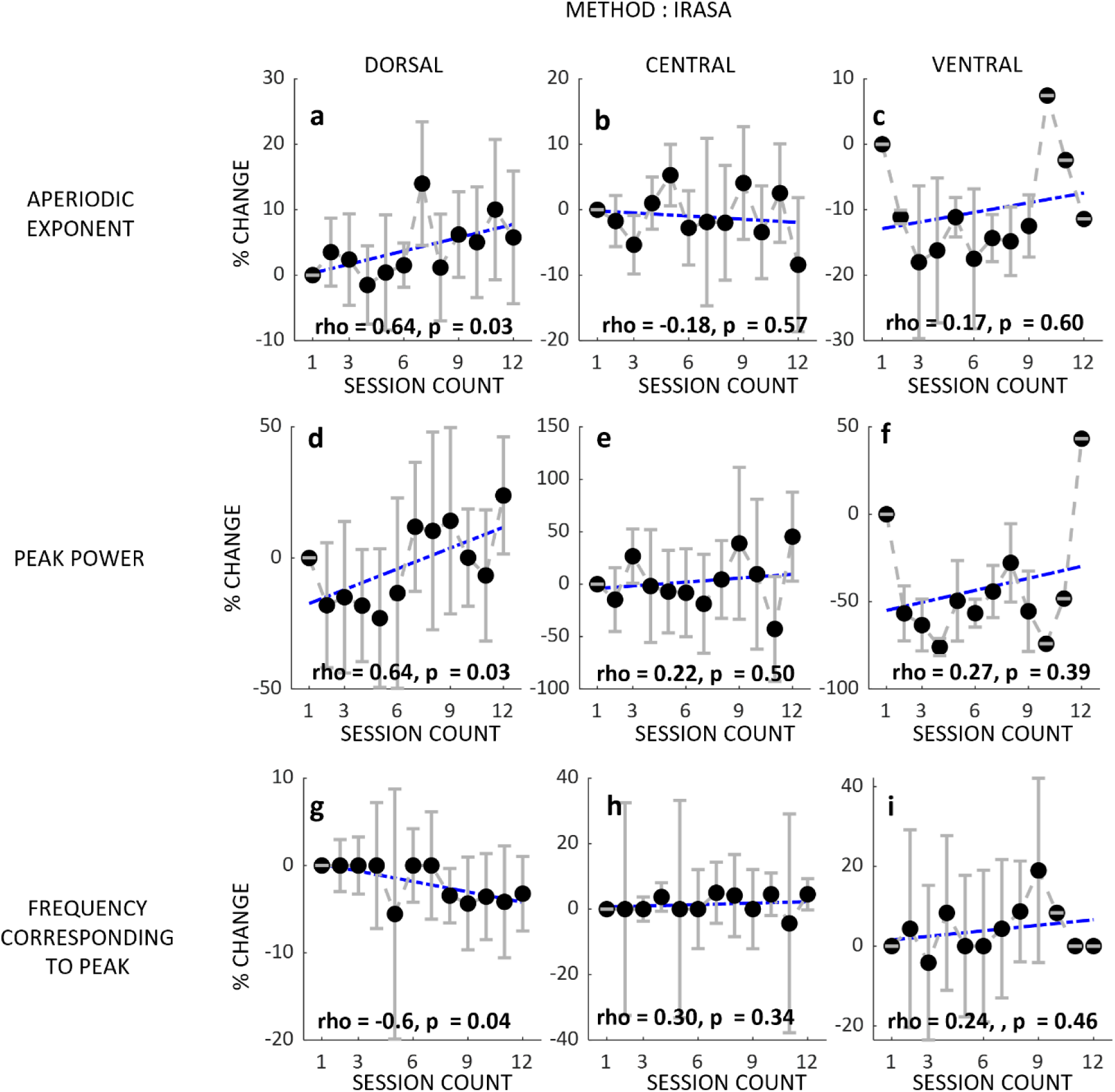
(a-i) illustrate the trend in the change in LFP features throughout the 12 exercise sessions. For each electrode placed dorsally, centrally, and ventrally, the percentage change from the respective baseline was calculated using the IRASA algorithm. The median value for all electrodes for each session is marked with a filled black circle, and the light grey error bars show the interquartile range. (a-c) depict the change in the aperiodic exponent in the dorsal, central, and ventral regions, respectively. (d-f) show the incremental changes in peak power detected within the 8-50 Hz frequency range, and (g-i) display the changes in the frequency corresponding to the peak values as the sessions progress.

### Changes in the motor scores

Changes in LFP activity also correlated with motor improvement. We found significant enhancements in postural tremor (66.76±32.78% improvement over 4 weeks) and kinetic tremor (40.29±22.17% improvement) in response to exercise, however, minimal resting tremor was observed even at the pre-exercise state since DBS was on. In other words, dynamic cycling improved postural and kinetic tremor that was refractory to DBS at programmed parameters. Finger taps speed and rhythm improved by 20±36% and 28.8±77.13%, respectively. Although hand movement speed remained unchanged, there was a notable improvement in hand movement rhythm by 32.83±30.57%. Dynamic cycling did not improve the foot taps.

## Discussion

Expanding on previous evidence of beneficial impact of dynamic cycling on PD motor symptoms (Gates 2022, Alberts 2016, Ridgel 2015, Ridgel 2009), this pilot study aimed to uncover the mechanisms behind these improvements. Our hypothesis centered on the modulation of STN activity measured by LFP in PD, both acutely and in the long term. Using DBS electrodes, we measured LFP frequency, power at the dominant spectral frequency, and aperiodic parameters (1/f exponent). We found no immediate conclusive effects of dynamic cycling; however, lasting, and incremental effects were evident over the 4-week intervention, with up to 12 sessions of dynamic cycling in the dorsal part of the STN. We were able derive the same conclusion from 2 different methods to identify the aperiodic spectrum of the LFP recordings. FOOOF and IRASA are two distinct and well received methods for calculating the aperiodic exponent and identifying the aperiodic component of the spectrum. Their strengths and weaknesses have been discussed thoroughly by (Gerster 2022). To avoid errors in interpreting the results due to the limitations of a single method, we conducted analyses using both and compared the results to validate our conclusions.

Both periodic and aperiodic activity, measured by the power at peak frequency and the 1/f exponent, can vary significantly and may be influenced by different states of the nervous system, including responses to levodopa medication. Ideally, to isolate the effects of exercise from changes in dopaminergic concentration, we would have preferred participants to refrain from taking their levodopa medication overnight. However, since this was also a long-term intervention, it was not feasible to ask participants to withhold their medication. Hence, the dopamine concentration would likely vary between the before and after exercise readings, as the ingested medication is absorbed or eliminated from the body. This variation can either enhance or mask changes in the LFP features, as levodopa medication is known to affect various aspects of LFP signals, such as peak power, aperiodic exponent, and beta burst dynamics (Tinkhauser et al., 2017). To minimize the influence of dopaminergic medications on our interpretations, we structured our experiments with a consistent interval between medication intake, exercise, and the measurement of LFPs before and after dynamic cycling.

The long-term analysis revealed a gradual response in the dorsal STN over the course of 12 sessions. While the immediate response in the STN was not pronounced, there were observable neurophysiological changes over time. The aperiodic exponent showed an increasing trend as in the dorsal part of STN as the sessions progressed. The aperiodic exponent, serving as a measure of aperiodic activity in the recorded LFP signal, can be considered a surrogate measure of ionic conductance and the extracellular medium (Destexhe et al., 1993). Additionally, it has been proposed that this exponent reflects the excitatory-inhibitory balance in the region (Gao et al., 2017; Voytek et al., 2015; Wiest et al., 2023) When a person takes levodopa medication, an increase in the aperiodic exponent is observed (Wiest 2023), suggesting that the medication and exercise’s effect on the aperiodic spectrum may follow same neuronal pathways. However, the long-term trend in peak power in response to exercise does not mimic its response to dopamine intake. In PD patients, levodopa consumption typically reduces peak power, indicating a suppression of hypersynchrony in the STN, which is often correlated with symptom severity. Our data indicates an increase in the hypersynchrony in the dorsal region of the STN. One possible explanation for this can be that the exercise engages different pathways compared to medication for controlling the oscillatory component of the data. This needs to be validated with more experiments involving different parts of the brain involved in motor control. Additionally, the third LFP feature, the frequency corresponding to the peak power, showed a decreasing trend, suggesting that hypersynchrony is reducing in frequency, which may indicate a slowing down of information flow in the STN. Although the magnitude of the frequency changes is small, they might exhibit a more pronounced decreasing trend over a longer intervention period. A timeframe of 4 weeks is relatively short for an exercise study. For patients with advanced PD, it may take more time for exercise interventions to show significant effects. However, this study suggests that consistent physical activity can start to modify the behavior of the dorsal part of the STN. Many studies reporting improvements in PD patients with exercise have conducted interventions spanning 6 months to many years (Bhalsing 2018, Mak 2017, Laat 2024, Tsukita 2022). Therefore, to fully understand the extent of improvements, changes, and limitations that exercise can offer, a longer-duration study with these measurements is recommended.

While some studies show immediate changes in cortical activity after exercise, deeper brain structures like the STN may take longer and more consistent interventions to induce neuroplasticity. The extended effects are aligned with the notion that exercise induces nervous system plasticity, and such plasticity then led to changes in STN activity manifest in LFP changes. There is strong evidence that exercise and physical activity in PD involves neurotrophic factors as key mediators of neuroplasticity(Bettio et al., 2019). Intensive treadmill training is known to rescue alterations in striatal plasticity in rodent models of synuculopathy(Marino et al., 2023). Improved motor control is also associated with a recovery in dendritic spine density and alterations and lasting rescue of physiological corticostriatal long-term potentiation. Further pharmacological analyses of long-term potentiation have shown modulation of N-methyl-d-aspartate receptors bearing GluN2B subunits and tropomyosin receptor kinase B, BDNF is also involved in these beneficial effects(Marino et al., 2023). The exercise induced alterations in both dopaminergic and glutamatergic neurotransmission are thought to mitigate cortically driven hyper-excitability in basal ganglia that is seen in PD(Petzinger et al., 2010).

We observed significant trends only in the dorsolateral part of the STN, with no significant trends in the central or ventral sections of the lateral STN. We had only three electrodes placed in the ventral region of the STN and therefore prefer not to draw conclusions from that data. However, there were around 11 electrodes placed in the central part of the lateral STN. The dorsolateral part of the STN, also known as the sensorimotor region, is connected to the motor cortex via the hyper direct pathway. The more ventral regions are connected with the limbic and associative circuits, which did not show a response to the exercise paradigms.

There are numerous factors contributing to the variability in the data. Despite the participant cohort comprising PD patients, variables such as fitness levels, regular exercise routines, age, and specific symptom presentations likely contribute to this variability. Additionally, individual differences in response rates to exercise regimes may also vary.

DBS is a powerful technique to mitigate PD symptoms. When the stimulation is withdrawn, motor symptoms may take up to 4 hours to fully return (Temperli 2003). Although the functional symptoms manifest gradually, the reemergence of beta peaks in the LFPs suppressed during stimulation can be observed in the data. By maintaining a consistent interval between turning off the stimulation and recording the data, we can still obtain reliable results. However, this approach prevents us from accurately measuring the change in motor symptoms due to dynamic cycling while DBS is turned on. When DBS is turned on, many symptoms are already suppressed, making it difficult to claim improvements. Nonetheless, we still observed some improvements in certain motor tasks. In otherwords, dynamic cycling was able to address motor symptoms that were refractory or suboptimal treated with STN DBS.

Throughout the month-long experiment, we do not expect significant shifts in the participants’ brain states due to disease progression. Considering that the reported mean annual decline for PD patients in the Hoehn and Yahr scale is approximately 3.2% per year(Alves et al., 2005), translating to less than 0.3% monthly, and our mean UPDRS-III score for these participants was 20, a 0.3% change in 20 amounts to a mere 0.06 alteration in the UPDRS score by assuming linear changes, indicating minimal disease progression within a month. Hence, it seemed reasonable to attribute any observed changes to the exercise intervention rather than disease progression.

In summary, this pilot study examines the impact of high cadence dynamic cycling on the STN activity in PD patients, investigating acute and long-term changes in LFPs. While immediate post-exercise effects were not observed, significant alterations in STN LFP power and periodicity emerged over a four-week, 12-session period for the dorsal region of the STN. These findings indicate dynamic cycling induces neuroplastic changes in STN activity, potentially enhancing motor function in PD patients. The study underscores the significance of prolonged exercise interventions for managing PD symptoms.

## Data Availability

Due to limitations from the IRB, the data is not available.

## Acknowledgements

Our gratitude extends to the Medtronic, Minneapolis, MN, USA team particularly James Adler and Robyn Whipple, and Adam Matthews for their invaluable support in steering us through the data collection and raw data interpretation process. We also extend our thanks to the whole team at InMotion, Beachwood, Ohio, USA for providing us the support and facility to conduct our experiments.

## Author’s Roles

P.J.: Study design, data collection, data analysis, writing-original draft.

L.M.F: data collection, writing - review and editing.

B.E.S: data collection, writing - review and editing.

C.W.K: participant recruitment, data collection, writing - review and editing.

A.G: Data analysis, editing.

K.A.L writing-review and editing.

A.L.R: project administrations and supervision, writing - review and editing.

A.G.S: Study design, data collection, participants recruitment, project administrations and supervision, Writing-original draft, review and editing

## Financial Disclosure and Conflict of Interest

Angela Ridgel and Kenneth Loparo are co-inventors on two patents which are related to the device used in this study: “Bike System for Use in Rehabilitation of a Patient,” US 10,058,736. No royalties have been distributed from this patent.

**Table 1:**
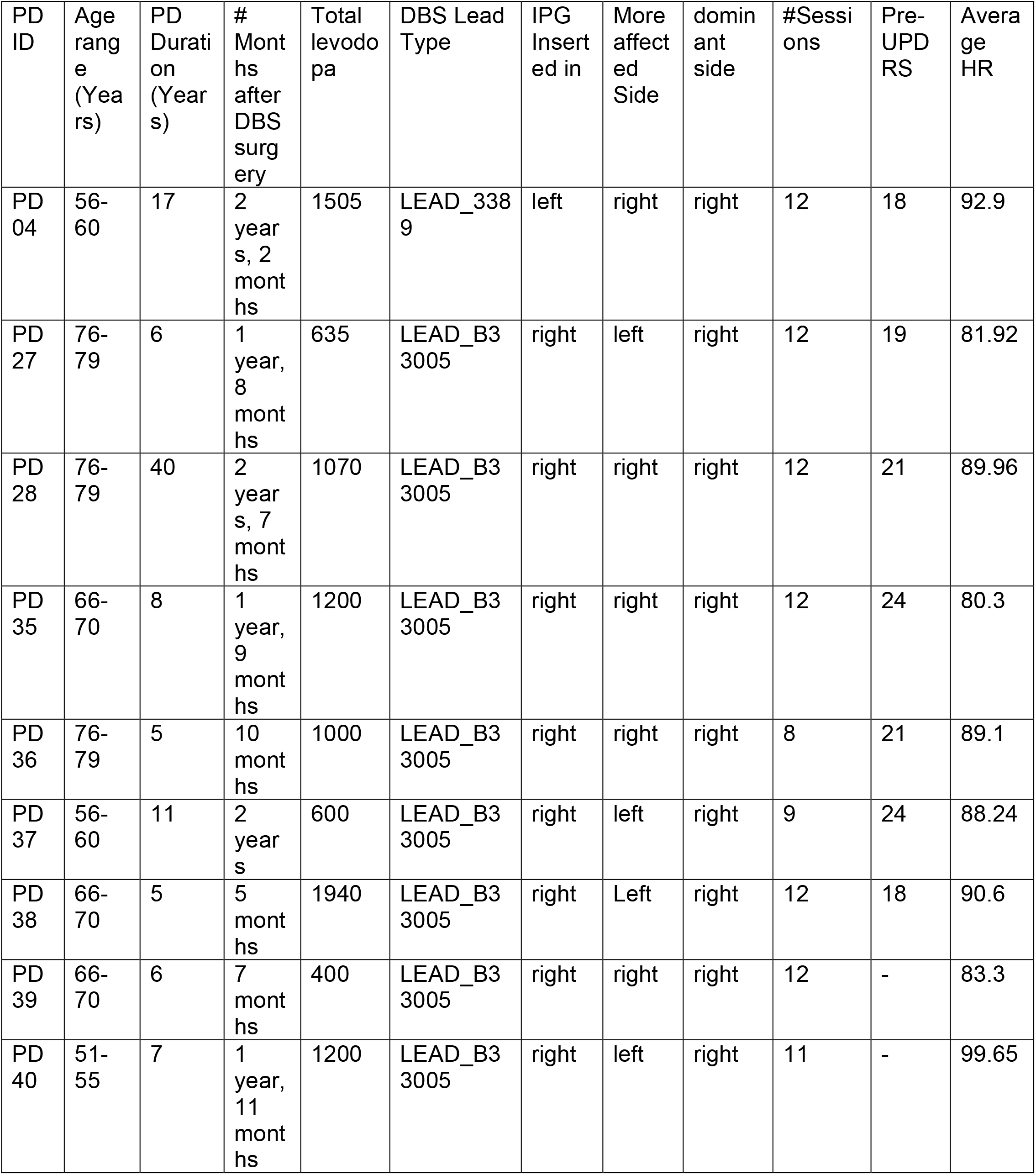
Highlights the demographic information about the participants, implantation, and their exercise summary. (IPG – Implanted Pulse Generator, UPDRS – Unified Parkinsons’s Disease Rating Scale, HR – Heart Rate). The participant IDs listed in the tables have not been shared with anyone outside the research group.

| PD ID | Age range (Years) | PD Duration (Years) | # Months after DBS surgery | Total levodopa | DBS Lead Type | IPG Inserted in | More affected Side | dominant side | #Sessions | Pre-UPDRS | Average HR |
| --- | --- | --- | --- | --- | --- | --- | --- | --- | --- | --- | --- |
| PD 04 | 56-60 | 17 | 2 years, 2 months | 1505 | LEAD_3389 | left | right | right | 12 | 18 | 92.9 |
| PD 27 | 76-79 | 6 | 1 year, 8 months | 635 | LEAD_B33005 | right | left | right | 12 | 19 | 81.92 |
| PD 28 | 76-79 | 40 | 2 years, 7 months | 1070 | LEAD_B33005 | right | right | right | 12 | 21 | 89.96 |
| PD 35 | 66-70 | 8 | 1 year, 9 months | 1200 | LEAD_B33005 | right | right | right | 12 | 24 | 80.3 |
| PD 36 | 76-79 | 5 | 10 months | 1000 | LEAD_B33005 | right | right | right | 8 | 21 | 89.1 |
| PD 37 | 56-60 | 11 | 2 years | 600 | LEAD_B33005 | right | left | right | 9 | 24 | 88.24 |
| PD 38 | 66-70 | 5 | 5 months | 1940 | LEAD_B33005 | right | Left | right | 12 | 18 | 90.6 |
| PD 39 | 66-70 | 6 | 7 months | 400 | LEAD_B33005 | right | right | right | 12 | - | 83.3 |
| PD 40 | 51-55 | 7 | 1 year, 11 months | 1200 | LEAD_B33005 | right | left | right | 11 | - | 99.65 |

**Table 2:**
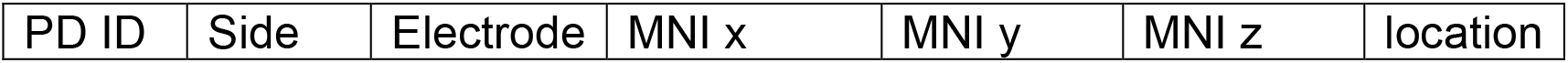
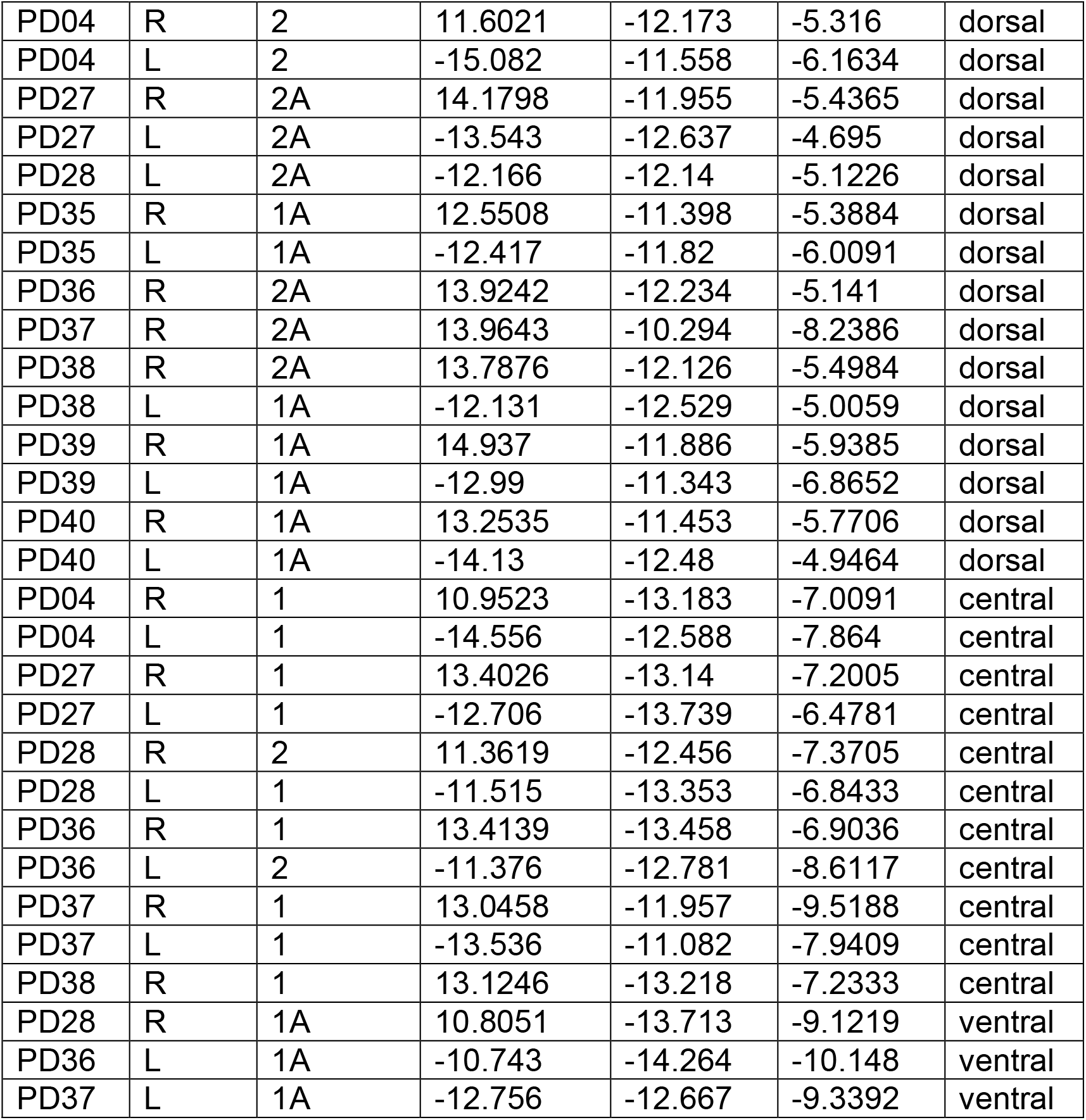
Displays the MNI (Montreal Neurological Institute) space co-ordinates of the electrodes of the electrodes used in the analysis from 9 participants. The participant IDs listed in the tables have not been shared with anyone outside the research group.

| PD ID | Side | Electrode | MNI x | MNI y | MNI z | location |
| --- | --- | --- | --- | --- | --- | --- |
| PD04 | R | 2 | 11.6021 | -12.173 | -5.316 | dorsal |
| PD04 | L | 2 | -15.082 | -11.558 | -6.1634 | dorsal |
| PD27 | R | 2A | 14.1798 | -11.955 | -5.4365 | dorsal |
| PD27 | L | 2A | -13.543 | -12.637 | -4.695 | dorsal |
| PD28 | L | 2A | -12.166 | -12.14 | -5.1226 | dorsal |
| PD35 | R | 1A | 12.5508 | -11.398 | -5.3884 | dorsal |
| PD35 | L | 1A | -12.417 | -11.82 | -6.0091 | dorsal |
| PD36 | R | 2A | 13.9242 | -12.234 | -5.141 | dorsal |
| PD37 | R | 2A | 13.9643 | -10.294 | -8.2386 | dorsal |
| PD38 | R | 2A | 13.7876 | -12.126 | -5.4984 | dorsal |
| PD38 | L | 1A | -12.131 | -12.529 | -5.0059 | dorsal |
| PD39 | R | 1A | 14.937 | -11.886 | -5.9385 | dorsal |
| PD39 | L | 1A | -12.99 | -11.343 | -6.8652 | dorsal |
| PD40 | R | 1A | 13.2535 | -11.453 | -5.7706 | dorsal |
| PD40 | L | 1A | -14.13 | -12.48 | -4.9464 | dorsal |
| PD04 | R | 1 | 10.9523 | -13.183 | -7.0091 | central |
| PD04 | L | 1 | -14.556 | -12.588 | -7.864 | central |
| PD27 | R | 1 | 13.4026 | -13.14 | -7.2005 | central |
| PD27 | L | 1 | -12.706 | -13.739 | -6.4781 | central |
| PD28 | R | 2 | 11.3619 | -12.456 | -7.3705 | central |
| PD28 | L | 1 | -11.515 | -13.353 | -6.8433 | central |
| PD36 | R | 1 | 13.4139 | -13.458 | -6.9036 | central |
| PD36 | L | 2 | -11.376 | -12.781 | -8.6117 | central |
| PD37 | R | 1 | 13.0458 | -11.957 | -9.5188 | central |
| PD37 | L | 1 | -13.536 | -11.082 | -7.9409 | central |
| PD38 | R | 1 | 13.1246 | -13.218 | -7.2333 | central |
| PD28 | R | 1A | 10.8051 | -13.713 | -9.1219 | ventral |
| PD36 | L | 1A | -10.743 | -14.264 | -10.148 | ventral |
| PD37 | L | 1A | -12.756 | -12.667 | -9.3392 | ventral |

